# Covid-19 pandemic has polarized the society toward negative and positive traits depending on a person’s resilience and certain predispositions

**DOI:** 10.1101/2022.10.25.22281466

**Authors:** Taraneh Attary, Leila Noorbala, Ali Ghazizadeh

## Abstract

The Covid-19 pandemic has caused a major disruption affecting almost all aspects of health, social and economic dimensions of our lives on an almost unprecedented global scale. While Covid-19 itself is, first and foremost, a pernicious physical illness, its highly contagious nature has caused significant psychological stress with occasional dire mental health consequences which are still not fully understood. To address this issue, we have conducted a longitudinal study by administering standard self-reporting questionnaires covering five major personalities and six mental traits of subjects before and a few months after the outbreak. Results revealed the distribution of population scores to become more extreme in either positive or negative trait directions despite the stability of average trait scores across the population. Higher resilience was found to be positively correlated with improved trait scores post-pandemic. Further investigations showed that certain predispositions could have an effect on trait score change post-Covid depending on the subject’s pre-Covid scores. In particular, in the subjects with moderate scores, there was a significant negative correlation between the positive trait scores and the post minus pre-positive trait scores. By examining various traits and personalities, these findings depict a more thorough picture of the pandemic’s impact on society’s psychological well-being and reveal certain predispositions and vulnerabilities that shape the mental health landscape in the post-Covid period with implications for mental health policies in dealing with Covid-19.

## Introduction

The spread of Covid-19 has caused unprecedented challenges for people worldwide, particularly due to the drastic degradation of social life and increased isolation. The pandemic has not only affected people’s physical health but also had a significant impact on people’s mental health probably due to chronic psychological stress (1–3). As the Covid-19 pandemic continues, it is predicted that we will face a long-term global mental health crisis (4–8). To plan for and remedy the resulting mental consequences, it is imperative to have a detailed and quantitative understanding of the psychological impacts of the Covid-19 pandemic across a wide range of traits and mental dispositions.

Previous work has addressed the effects of Covid-19 on mental disorders such as depression, anxiety (3,9–12), and post-traumatic stress disorder (PTSD) (13,14). Moreover, there has been research on the role of people’s personalities and dispositions in the Covid pandemic such as neuroticism, extraversion, openness, agreeableness, conscientiousness (15,16), alexithymia (17–19) and autistic characteristic (6,20,21). However, to-date, no studies have considered the effect of the Covid-19 simultaneously on multiple traits and personalities in the same population before and after the pandemic. In particular, considering a cohort of traits in a within-subject design is desirable for gaining a much more accurate and complete picture of the pandemic consequences and the possible heterogeneities in the affected population.

Here, we report the results of a longitudinal study involving 114 subjects that participated in a study to answer seven standard questionnaires which included the NEO-five factor inventory (NEO-FFI), shyness, alexithymia, autism quotient, anxiety, depression, and sensory processing sensitivity (SPS) covering a total of 11 traits a few months before (Materials and Methods) and again a few months after the Covid pandemic (pre- and post-pandemic periods, respectively). These subjects also completed the mental resilience and Corona anxiety questionnaire during the pandemic to examine their possible roles in the mental health state in the post-pandemic period. As expected, results show that individuals with higher resilience were less negatively impacted by the pandemic. Interestingly, certain predispositions also predicted their response to the pandemic in the post-era.

## Results

To address how the Covid-19 pandemic might have affected the individual personalities and traits, we compared the subjects’ scores (n=114) to questionnaires spanning 11-traits before the start of the pandemic and about four months after the pandemic (pre and post-Covid periods, respectively). Consistent with our previous report (22), the traits were found to be grouped in two main clusters by unsupervised hierarchical clustering (HC). The first trait cluster which included openness, extraversion, conscientiousness, and agreeableness constituted the positive trait cluster. The second trait cluster, which included autism, shyness, alexithymia, anxiety, depression, neuroticism, and sensory processing sensitivity formed the negative trait cluster. Notably, both trait clusters were stably found in pre and post-Covid periods (Fig 1a-b).

**Figure 1:**
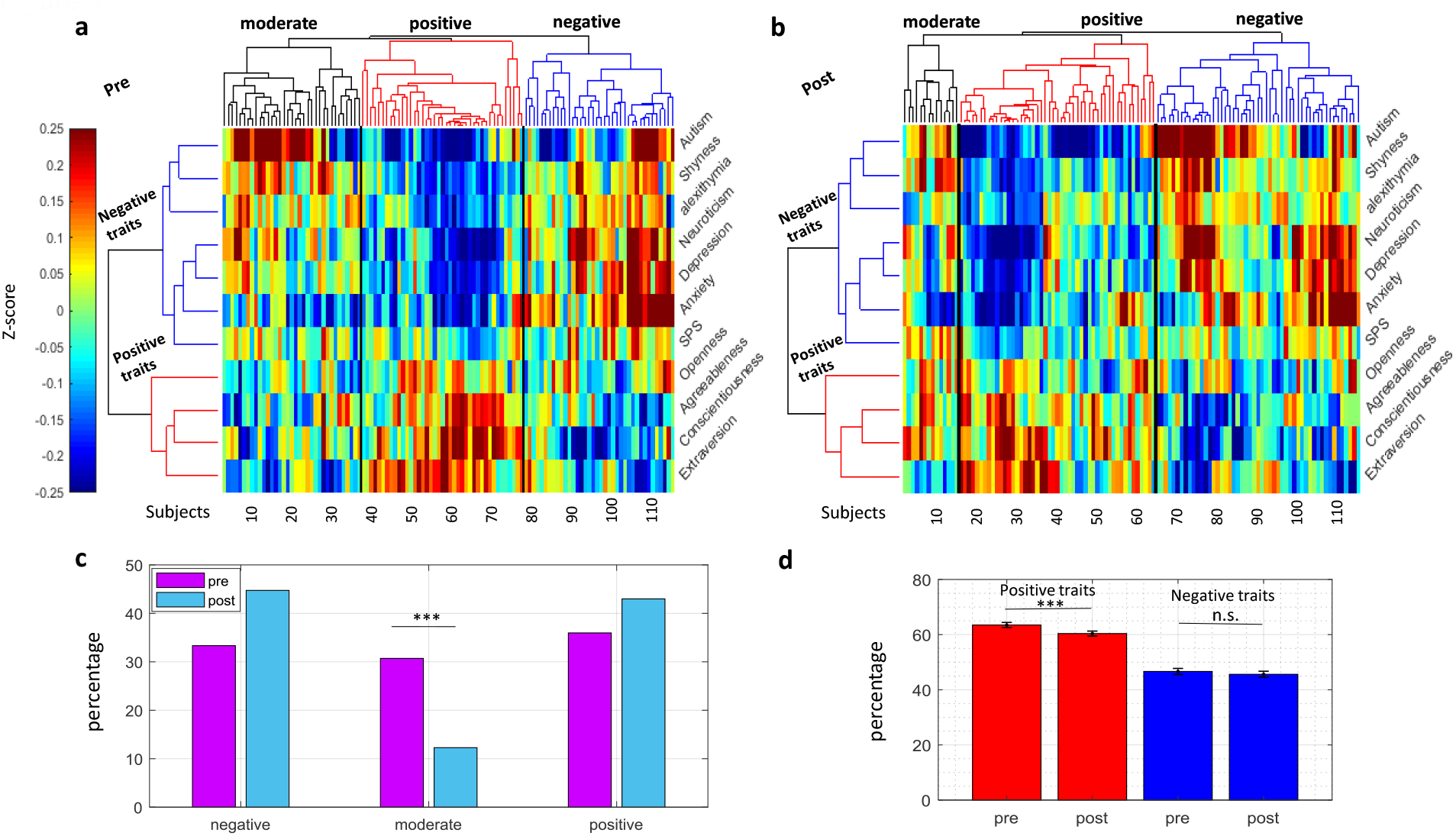
Change of subject groups before and after Covid-19. Clustergram of subjects (columns) before Covid-19 (a) and after Covid-19 (b) color-coded by z-scores of 11 traits (rows) with rows and columns sorted by HC. c, Percent of subjects in the three subject groups before and after Covid-19. d, Average score of positive traits and negative traits clusters before Covid-19 (t_113_= 9.23) and after Covid-19 (t_113_=8.19), ***p<1e-3. Pre= before Covid-19, post = after Covid-19. Error bars represent s.e.m. here and thereafter.

Subjects also showed three main groups using HC in both pre and post-Covid periods. These groups included subjects with high scores in positive traits and low scores in negative traits (positive subject group), subjects with high scores in negative traits and low scores in positive traits (negative subject group), and subjects with moderate negative and positive scores (moderate group) (Fig S1a-b). Interestingly, we found a significant shrinkage in the number of moderate subjects in the post compared to the pre-Covid period (Fig 1c). However, the subjects migrating out of the moderate group did not all transfer to the negative group as might be expected from the hardship inflicted by Covid. Notably subjects migrating out of positive and negative groups mostly did not end up in the moderate group but moved to the opposite extreme (Table 1). This resulted in a concurrent increase in the percentages of subjects in both positive and negative groups. This concurrent increase in positive and negative groups somewhat cancelled out each other such that the overall negative and positive population scores remained relatively stable in the post compared to the pre-Covid period (Fig 1d). The overall score of negative traits did not show a significant change from pre-Covid to post-Covid era (see figure captions for stats throughout). The positive trait scores showed only a small (3.2%) but significant reduction (Fig 1d). Thus, Covid-19 seems to have polarized the society into positive and negative groups while keeping the average positive and negative trait scores relatively stable across the population.

**Table 1:** Confusion matrix changes in the three subject groups before and after Covid-19. Confusion matrix showing the percentage of subjects in positive, moderate, and negative groups before (rows) and after (columns) the Covid-19 pandemic. The percentages are color coded and sum up to 100.

| pre/post (%) | positive | moderate | negative | Ppre(pre) |
| --- | --- | --- | --- | --- |
| positive | 27.19 | 2.63 | 6.14 | 35.96 |
| moderate | 7.01 | 8.78 | 14.92 | 30.71 |
| negative | 8.78 | 0.87 | 23.68 | 33.33 |
| Ppost(post) | 42.98 | 12.28 | 44.74 | 100 |

To address the factors that might have caused the differential migration of subjects toward positive and negative traits during the pandemic, we administered two additional questionnaires in the post Covid era for the subjects one measuring mental resilience (23,24) and the other measuring the degree of anxiety caused by the Covid pandemic (Corona anxiety, (25)). The resilience questionnaire consists of 25 questions and assesses the individual’s innate ability to deal with stressful environmental conditions (24) while the Corona anxiety questionnaire consists of 18 questions and assesses the degree of anxiety caused by Covid-19 (25). Across subjects, resilience and Corona anxiety showed a significant negative correlation (r= −0.19, p-value =0.04 Fig S1c). This suggests that subjects with higher resilience tended to experience less Corona anxiety and vice versa (without implying the causative direction). Furthermore, results showed that if added to the previous 11traits, resilience itself falls within the positive traits while Corona anxiety falls within the negative traits (Fig S1d).

We hypothesized that the subjects with higher resilience fared better during the pandemic and migrated to the positive group. In comparison, the subjects with lower resilience were more frequently found in the negative group. Indeed, results showed that the resilience score showed a positive correlation trend with the increase of subjects’ scores for positive traits from pre to post-Covid period (Pearson’s r= 0.16, p-value= 0.07) and was significantly and negatively correlated with the increase of subjects scores for negative traits going from pre to post-Covid period (Pearson’s r=-0.20, p-value=0.02) (Fig 2a-b). Consistently, further analysis showed that the subjects in the highest resilience quartile became almost exclusively concentrated in the positive group in the post-Covid era (93% post vs 75.2% pre in the positive group). The opposite trend was observed in the lowest resilience quartile where the subjects became heavily biased toward the negative group in the post-Covid era. (82.9% post vs 41.4% pre in the negative group) (Fig 2c). Interestingly, no significant correlation between Corona anxiety and change of positive/negative trait scores were observed suggesting that anxiety about Covid-19 by itself was not a key determinant of how subjects fared during the pandemic (Fig 2d-f). Additional analysis using logistic regression was performed to see which one of the Corona anxiety or resilience had better predictive power for the change of the subject groups. Consistently, results showed that resilience had a much larger effect on the change of the subject groups from pre to the post-covid era (coefficient of resilience and Corona anxiety are 0.24 (p-value=1e-6) and −0.04 (p-value=0.03), respectively).

**Figure 2:**
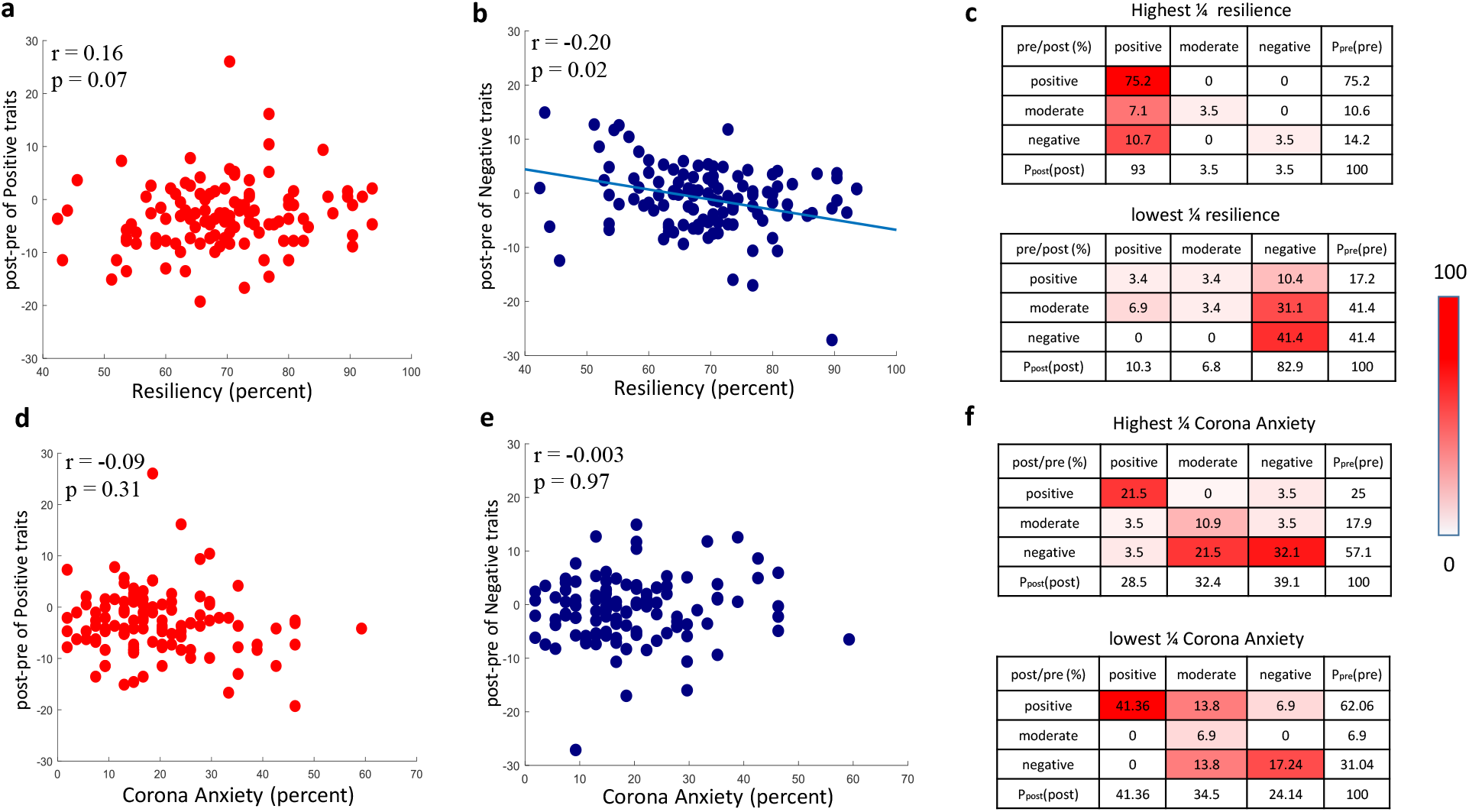
Change of post minus pre trait scores as a function of resiliency and Corona anxiety. a, Scatter plot of positive traits score change as function of resiliency across subjects. b, Same as a but for negative trait score change. c, Confusion matrixes showing the percent of subjects in positive, moderate, and negative groups before (rows) and after Covid-19 (columns) in subjects with highest resilience quartile, scores=76.8-93.6%, N=28, and lowest resilience quartile, scores=42.4-62.4%, N=29, respectively. d-e, Same format as a-b but for Corona anxiety for positive trait score change and negative trait score change. f, Same format as c but for highest Corona anxiety quartile, scores=25.9-85.18%, N=28, and lowest. Corona anxiety quartile, scores=1.8-12.9%, N=29. The best Deming regression line in the scatter plots is derived using the Deming method and it is shown only in the scatters where its p-value was significant in this figure and hereafter.

We repeated the correlations between resilience and post minus pre scores separately for positive, moderate, and negative subject groups in the pre-Covid era. Results showed that the resilience score was positively correlated with the increase of subjects’ scores for positive traits and negatively correlated with the increase of subjects’ scores for negative traits going from the pre to post-Covid period in all three subject groups (Fig 3a-b). Once again, Corona anxiety did not show a significant correlation with changes in trait score within any of the three subject groups’ consistent lack of an effect across all subjects (Fig S2a-b).

**Figure 3:**
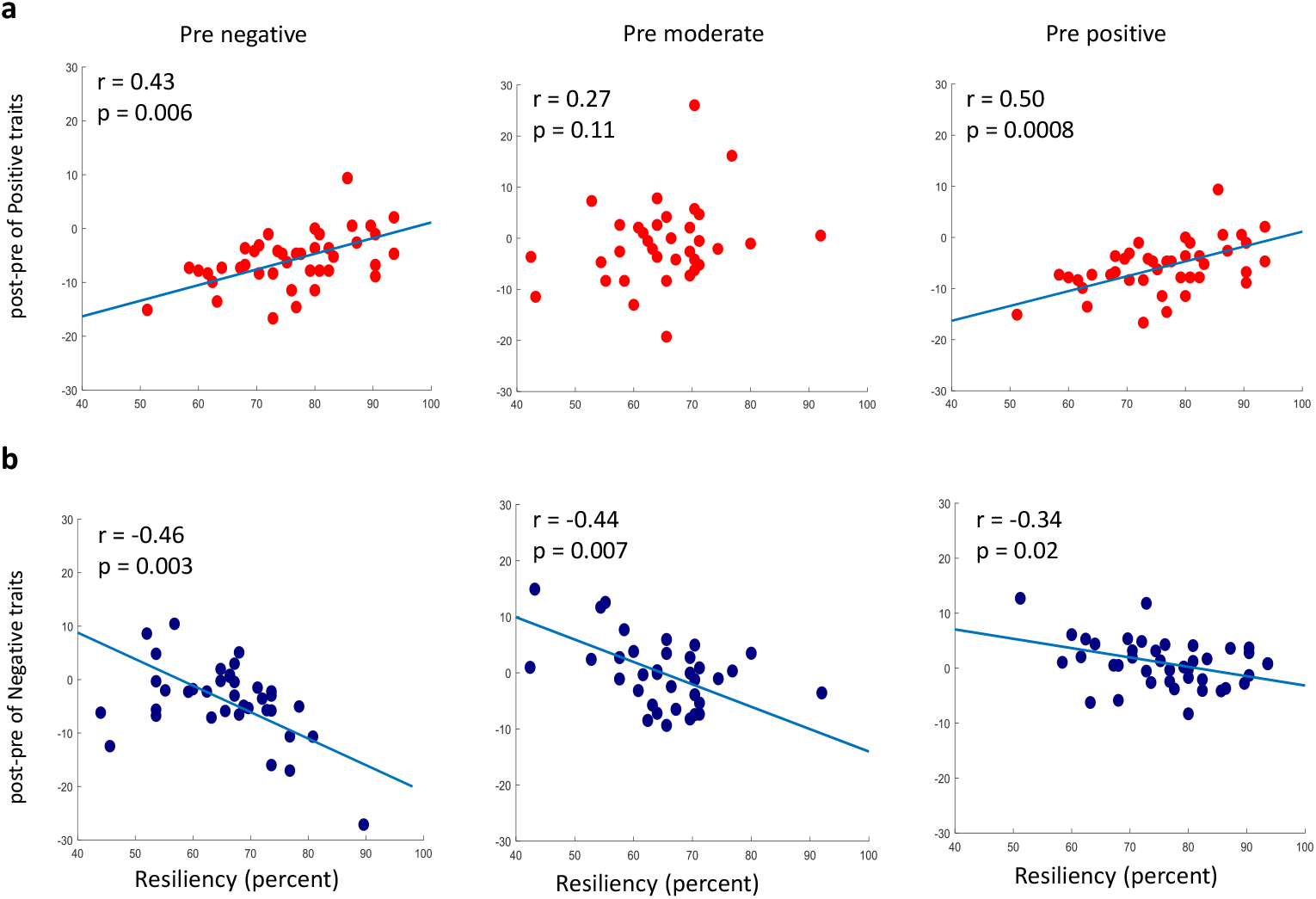
Correlation between resilience and trait score change. a, Scatter plot of positive trait score change and resilience separately in the three subjects groups. b, Same as a but for negative trait score change.

To investigate whether the subjects’ trait predispositions also played a role in the trait scores in the post-Covid era, the correlation between pre-Covid positive and negative trait scores with post minus pre scores were examined. For subjects in the negative or positive groups in the pre-Covid era, results showed almost no correlation between overall pre-Covid positive and negative train scores and the change in the trait scores (Fig 4a,c). However, for the subjects in the moderate group in the pre-Covid era, we found a significant negative correlation between the positive trait scores and the post minus pre-positive trait scores (r=-0.48, p-value=0.003) (Fig 4b). This means that in the moderate group, the subjects with a higher positive trait score in the pre-Covid era ended up with a larger reduction of their positive trait score in the post-Covid era. Additional analysis done separately for individual positive trait scores showed a significant negative correlation for agreeableness score (r=-0.36, p-value=0.03) (Fig S3a) and a negative trend for openness (r=-0.24, p-value=0.16), extraversion (r=-0.15, p-value=0.37) and conscientiousness (r=-0.26, p-value=0.12). These findings suggest that individuals with higher positive scores, in particular, agreeableness tend to be more adversely affected by the pandemic. There was not a significant correlation between pre-Covid negative scores, and positive trait score change in the moderate group (r=0.11, p-value =0.52); looking at individual negative trait scores showed a borderline positive correlation between autism and improvement in the post minus pre-positive trait scores (r=0.32, p-value =0.05, Fig S3b).

**Figure 4:**
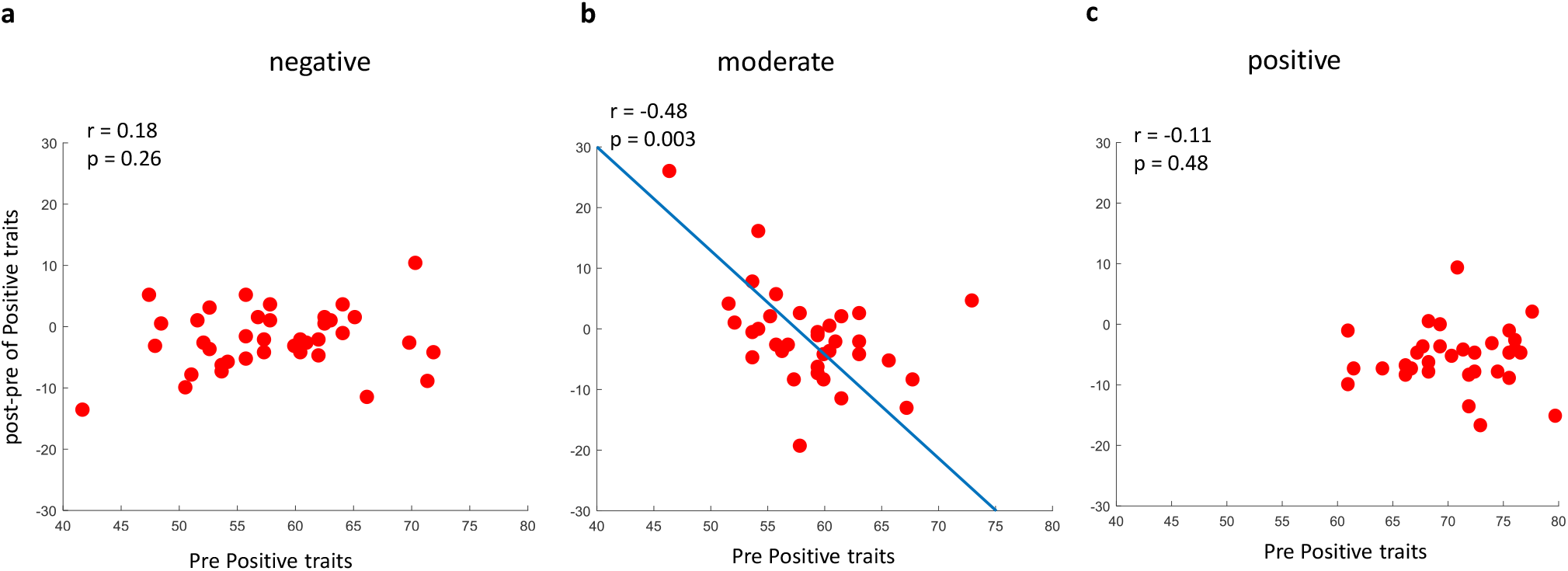
Correlation of positive trait score change and positive traits separately in a, negative b, moderate and c, positive subject groups

While there was no significant correlation between the overall pre-Covid scores in the negative group, some individual traits showed a significant correlation. There was a significant reduction of the post minus pre-negative scores for individuals with higher openness (r=-0.43, p-value=0.006) and for individuals with higher SPS scores (r=-0.37, p-value=0.02) (Fig S4a-b). There was also a borderline trend for a decrease of post minus pre-positive scores for individuals with higher shyness scores (r=-0.31, p-value=0.05) (Fig S4c). We found no effect of individual traits in individuals in the positive group (−0.28<r<0.16, p-value>0.07). However, we note if the p-values reported for individual traits are to be corrected using the most conservative family wise correction (e.g., Bonferroni), none of the individual trait effects remain significant. Nevertheless, the negative correlation between the positive trait scores and the post minus pre-positive trait scores remains significant even with Bonferroni correction (12 comparisons, corrected p-value=0.036, Fig S5)

## Discussion

Here, we report the results of a longitudinal study which tracked changes in 11 trait scores for subjects before and a few months into the Covid pandemic. The traits considered were divided into positive (openness, extraversion, conscientiousness, and agreeableness) and negative (autism, shyness, alexithymia, anxiety, depression, neuroticism, and sensory processing sensitivity) groups by unsupervised learning (22). Surprisingly, results showed that while Covid-19 significantly reduced the overall scores for positive traits, the effect was relatively small, and there was no significant change on the overall negative scores (Fig 1d). Instead, we found migration of subjects, and in particular ones with moderate scores to groups with more extreme positive and negative trait scores. We found two underlying factors correlated with the fate of subjects in the post Covid era. First, higher resilience (23) generally resulted in increased positive trait scores and decreased negative trait scores across the population (Fig 2). Unlike resilience, we did not find any significant effect on the degree of Corona anxiety. The second factor which specifically affected subjects with moderate trait scores, was their positive predispositions in the pre-Covid era such that higher positive scores in this group predicted a larger reduction of positive trait scores in the post-Covid era (Fig 4).

While the outbreak of the Covid-19 pandemic has certainly harmed the mental health of individuals, these effects have been relatively short-lived as people learned to adjust to the new social norms. In fact, reports show that as early as mid-2020, mental health conditions became almost comparable to the pre Covid period (26,27). In this regard, our post Covid score were collected about four months following the outbreak of the Covid 19 showed relatively small changes in the overall positive and negative trait scores, confirming previous findings. Despite this apparent stability in the overall scores, our results showed that Covid pandemic created a more bimodal distribution toward positive and negative groups and reduced the number of subjects in the moderate score group (Fig 1c). We found that the response of individuals to the pandemic was largely determined by their resilience scores. This confirm the important role of resilience in Covid pandemic, which in line with previous researches (28–31).

Interestingly, moderate subjects with higher agreeableness tended to have lower positive trait scores in the post-Covid era. Suggesting that social isolation for highly agreeableness people might have had a more detrimental effect. Consistently, previous research showed individuals with low agreeableness to be better equipped to adjust psychologically to problems imposed by Covid (31). The positive correlation between the individuals’ autism score and the improvement in the positive trait scores in the post-Covid era can also be interesting (Fig 4b). Indeed while the impact of Covid-19 is reported to be more severe for people with certain pre-existing mental problems (32–35), some improvements are also observed in people with attention deficit hyperactivity disorder (ADHD), bipolar disorder, and autism (36–40). One possibility is that social isolation during the pandemic era might have helped individuals with higher autism scores (39,40). This result emphasizes the importance of the big five in such phenomena (16,41). Unlike the general effects of resilience, these effects were mostly seen in subjects in the moderate group in the pre-Covid period.

In individuals in the negative group, higher openness scores resulted in lower negative scores post-Covid, while higher shyness scores resulted in lower positive scores (Fig S4). These results are consistent with previous literature that suggest openness to predict lower perceived stress and is positively associated with subjective well-being (31), whereas shyness is associated with negative feelings and for shy people being more fragile to psychopathology based on a lack of social interaction (42,43). What seems novel here is that these two traits predominantly exert their influence in the negative group, not the moderate or positive groups. Also, higher SPS had a positive effect in this group and was correlated with lower negative scores post-Covid. This is consistent with a previous finding that showed SPS not necessarily be a vulnerability factor in covid pandemic (44).

In summary, our results revealed that Covid-19 polarized society toward more extreme positive and negative trait scores with the heterogeneous response across the population depending on their resilience and certain predisposition in the few months following Covid-19. These individual score changes happened despite the overall stability of the average of positive and negative groups across the population. In particular the migration of the moderate subject group to more negative and positive traits and its dependence on the autism and agreeability scores, seems interesting and suggests that more social individuals may have felt the negative impacts of the pandemic and the subsequent social isolation more. Note that this research has tracked the individuals for a few months after the covid outbreak and further investigations are still needed as the pandemic continues globally. These findings may provide important clues for mental health policies for detecting and dealing with vulnerable groups in the post Covid era.

## Data Availability

All data produced in the present study are available upon reasonable request to the authors

## Acknowledgements

This research was supported by the Research Program at the Sharif University of Technology (Grant #G980703) and intramural grant at School of Cognitive Science, IPM.

## Author Contributions

TA, LN and AG conceived of and planned the study details. TA and LA collected data and TA analyzed the data under AG supervision. TA and AG wrote the paper with inputs from LN.

## Materials and Methods

### Subjects

From 837 subjects in our previous work (22) (prior to January 2020, It should be noted that the pandemic almost started in February in Iran), 114 subjects (71.93% female) with a mean age of 30.29 (SD = 11.04) filled out questionnaires again after few month into the pandemic (within June to September 2020 Period) voluntarily. These subjects also filled out mental resilience and Corona anxiety questionnaires.

### Questionnaires

The Persian version of seven questionnaires including Adult Autism Spectrum Quotient (45), Revised Cheek–Buss Shyness Scale (46), Toronto Alexithymia Scale (47), Beck Anxiety Inventory (48), Goldberg’s Depression scale (49), NEO Five-Factor Inventory (neuroticism, openness, extraversion, conscientiousness, and agreeableness) (50) and Highly Sensitive Person Scale (sensory processing sensitivity) (51) are used and described in the previous work (22).

Mental Resilience questionnaire is a self-report measure with 25-item (23). The response scale ranges from 0 to 5, from strongly disagree to strongly agree, respectively. The Alpha Cronbach of translated resilience questionnaire was 0.91 in this study.

Corona Disease Anxiety Scale (CDAS) questionnaire is an 18-item questionnaire to assess anxiety of people about Covid-19 (25). This questionnaire scale ranged from 0 to 3 never to always, respectively. The Alpha Cronbach of Persian questionnaire was 0.91 in this study.

### Data analysis

All data analyses were done by MATLAB 2017b. The scores for each question were z-scored across subjects before and during Covid pandemic separately for hierarchical clustering. For other analysis, the scores for each questionnaire were converted to a percentage. Logistic regression was done using MATLAB mnrfit. Hierarchical clustering and Alpha Cronbach calculation was done similar to our previous work (22). For Deming regression, equations are from York’s article in 1966 (52,53).

---

**Supplementary Figure 1:**
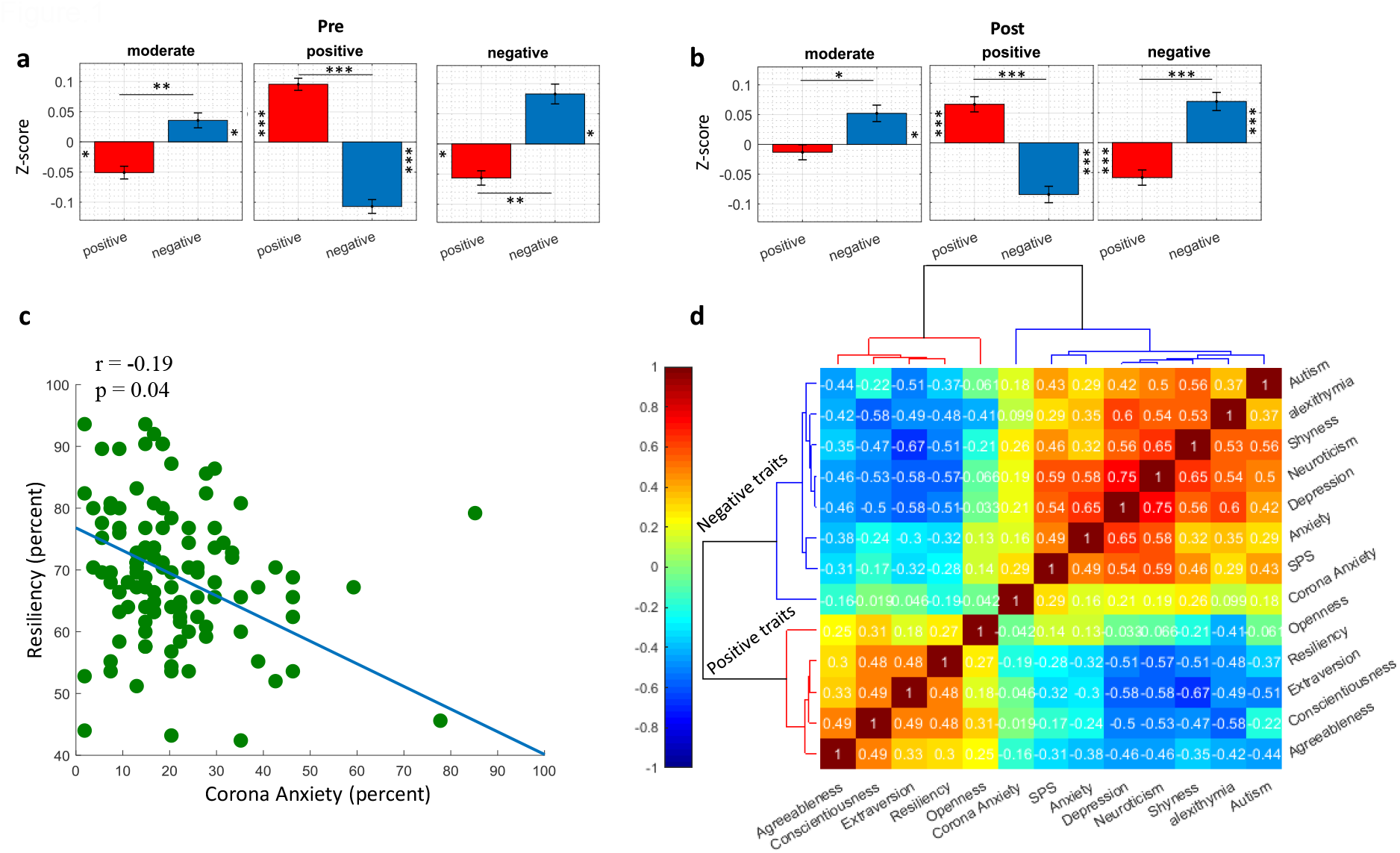
a, Average z-score of positive traits and negative traits cluster before Covid-19 in the three subject clusters of positive (t_40_ = 6.77), moderate (t_34_ = − 3.39), negative (t_37_ = − 3.50). b, Average z-score of positive traits and negative traits cluster after Covid-19 in the three subject clusters of positive (t_48_ = 7.13), moderate (t_13_ = − 2.12), negative (t_48_ = − 2.7). c, correlation of subject resiliency versus Corona anxiety. d, Color-coded pairwise correlation matrix of all traits during Covid-19 sorted by HC.

**Supplementary Figure 2:**
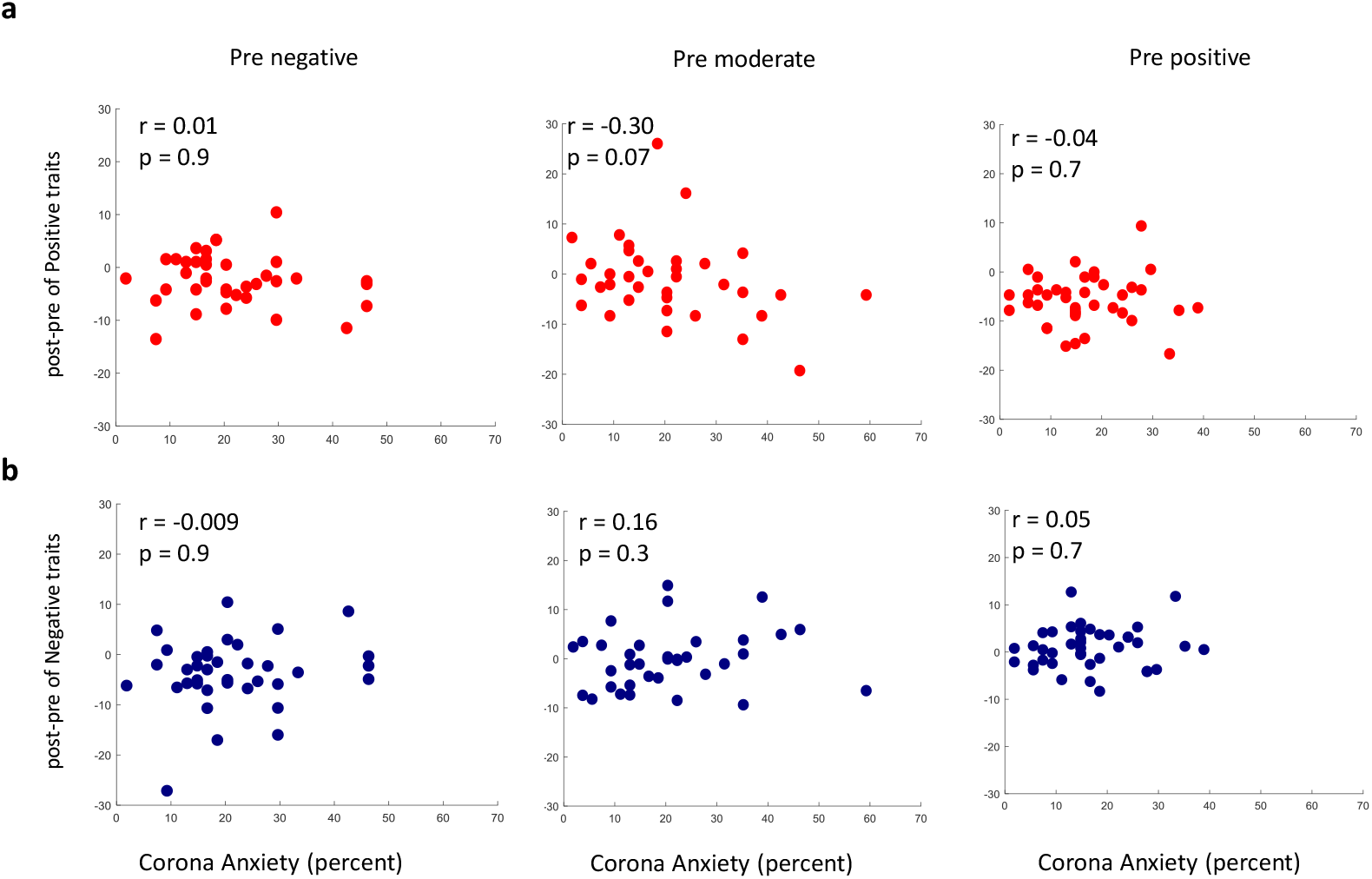
Same format as Figure 3 but for Corona anxiety.

**Supplementary Figure 3:**
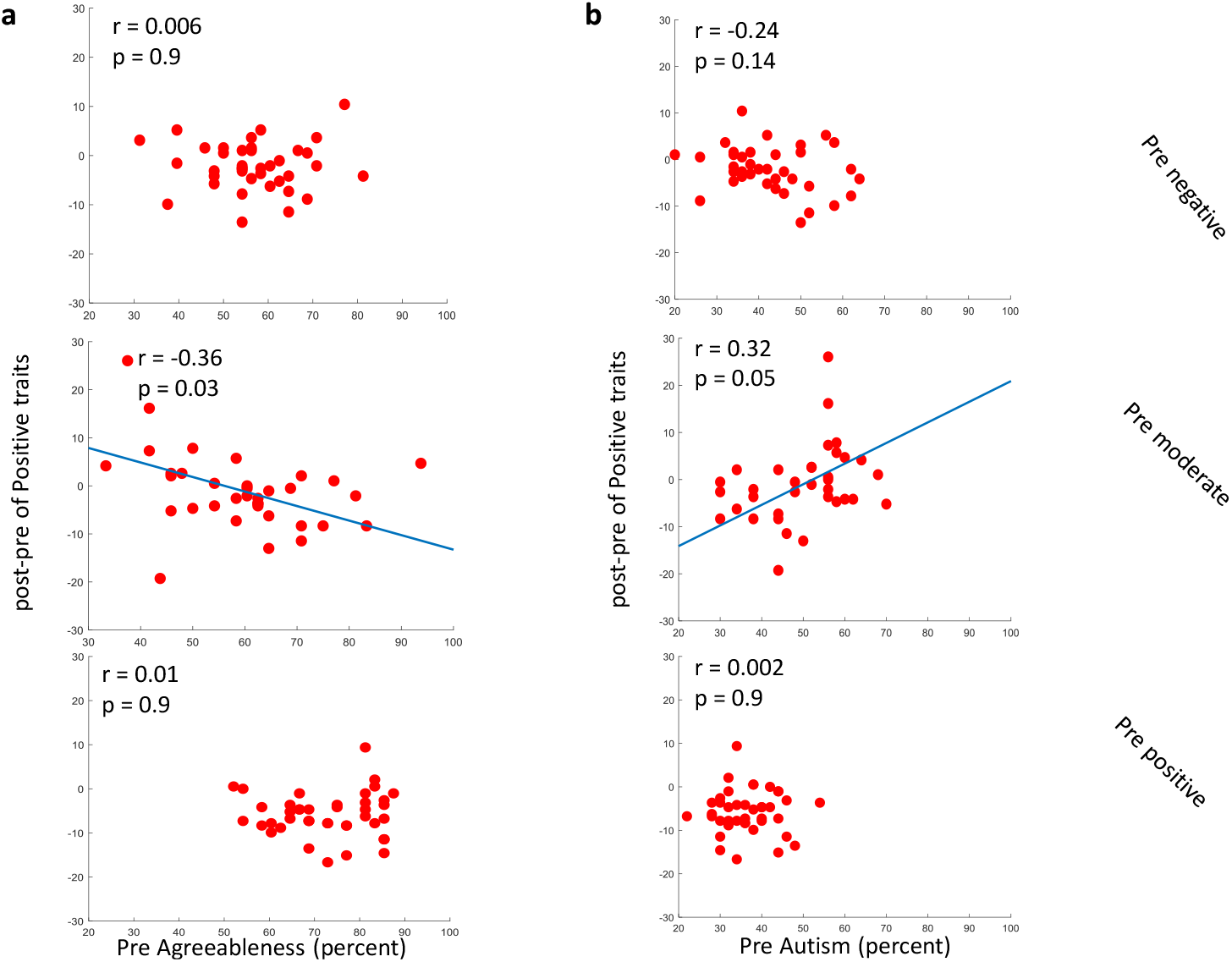
a, Scatter plot of positive trait score change versus agreeableness in the pre-Covid. b, Same as a but for autism in the pre-Covid era.

**Supplementary Figure 4:**
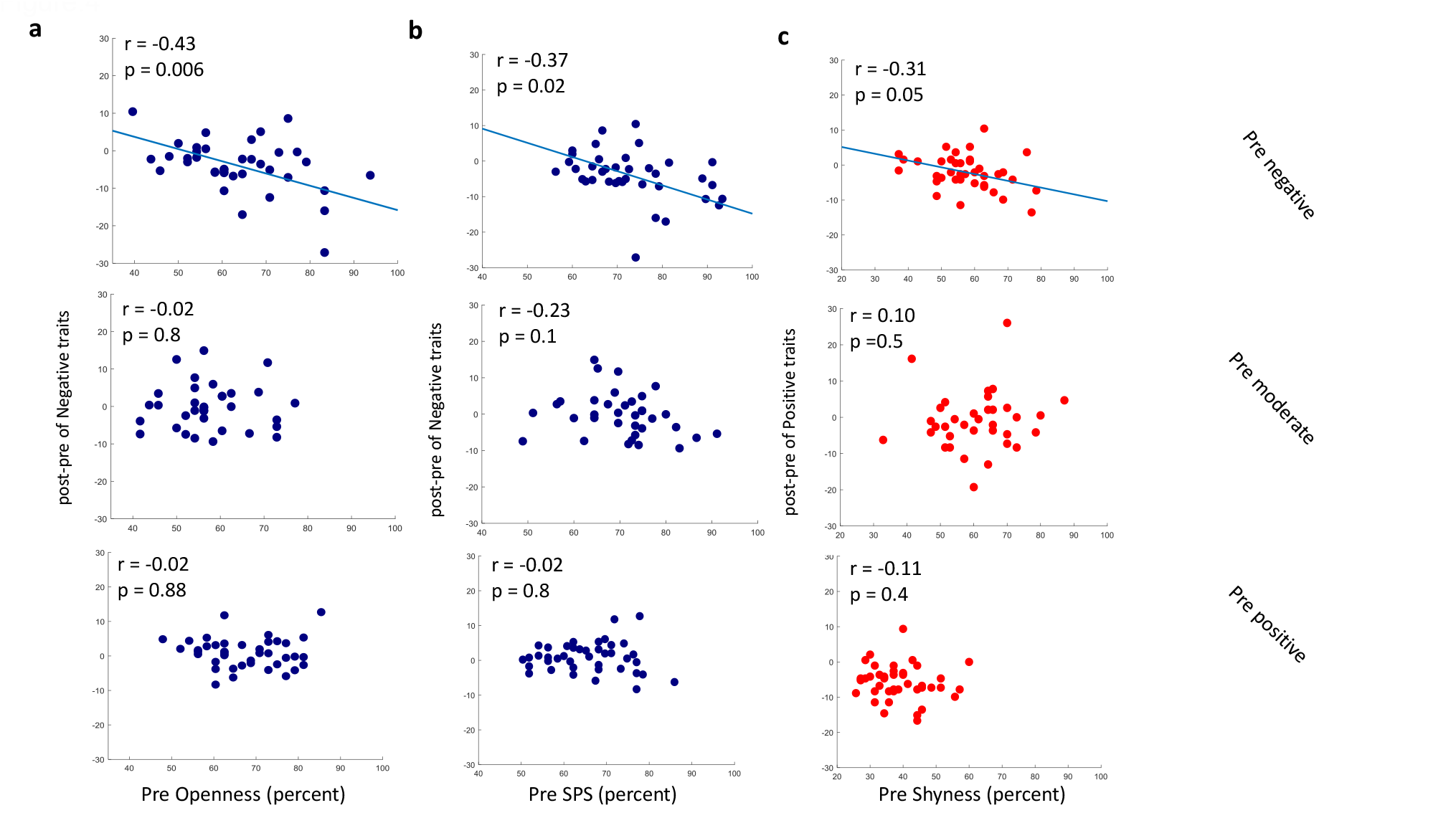
a, Scatter plot of negative trait score change versus openness in the pre-Covid era. b, Same as a but for SPS in the pre-Covid. c, Scatter plot of positive traits score change versus shyness in the pre-Covid era.

**Supplementary Figure 5:**
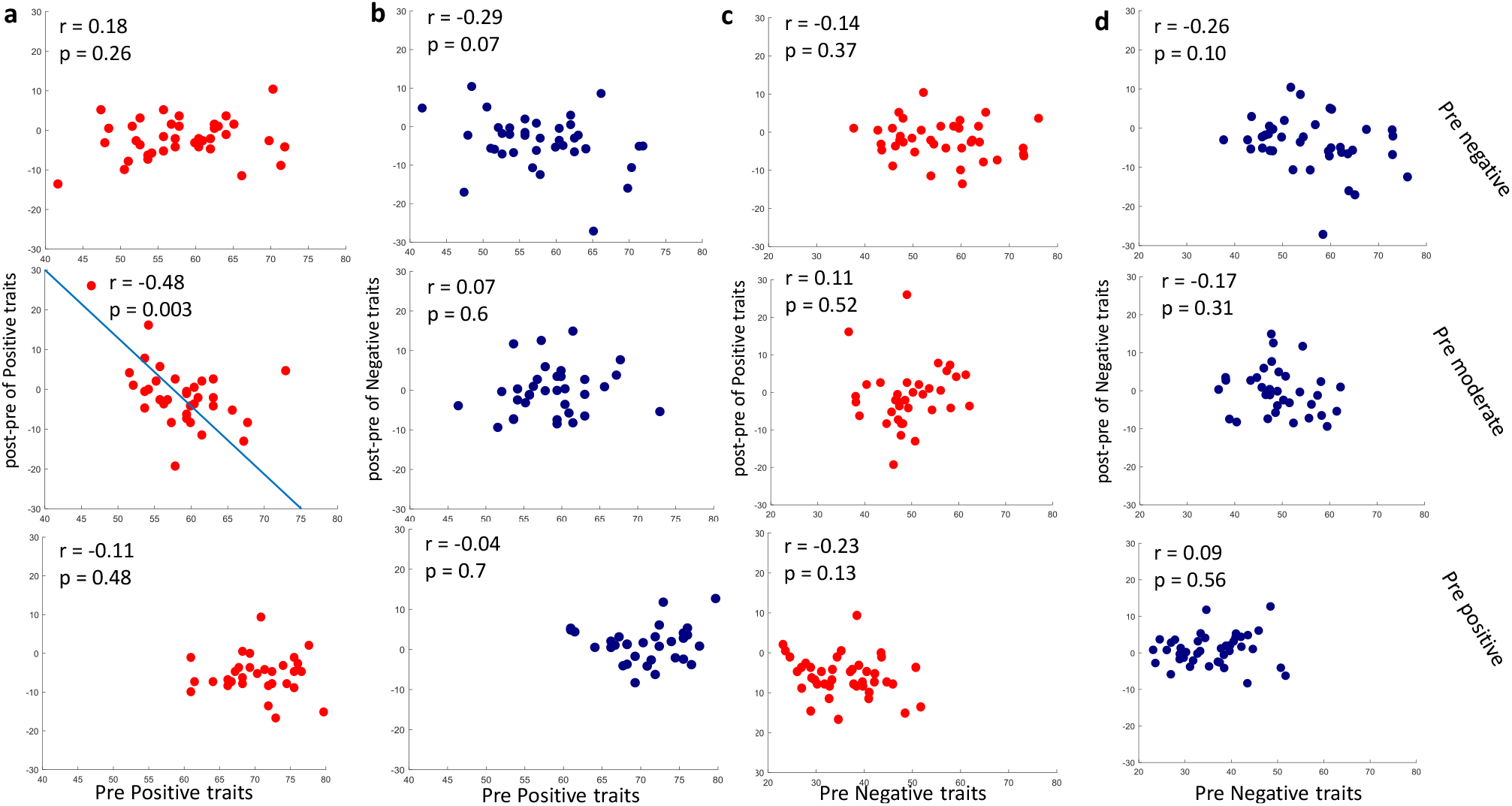
Trait score changes vs traits scores in the pre-Covid era. a, Scatter plot of positive trait score change and pre-Covid positive traits. b, Scatter plot of negative trait score change and pre-Covid positive traits. c, Scatter plot of positive trait score change and pre-Covid negative traits. d, Scatter plot of negative trait score change and pre-Covid negative traits.

